# Tuberculosis severity associates with variants and eQTLs related to vascular biology and infection-induced inflammation

**DOI:** 10.1101/2022.08.23.22279140

**Authors:** Michael L McHenry, Jason Simmons, Hyejeong Hong, LaShaunda L Malone, Harriet Mayanja-Kizza, William S. Bush, W. Henry Boom, Thomas R Hawn, Scott M. Williams, Catherine M. Stein

## Abstract

**Background:** Tuberculosis (TB) remains a major public health problem globally, even compared to COVID-19. Genome-wide studies have failed to discover genes that explain a large proportion of genetic risk for adult pulmonary TB, and even fewer have examined genetic factors underlying TB severity, an intermediate trait impacting disease experience, quality of life, and risk of mortality. No prior severity analyses used a genome-wide approach.

**Methods and Findings:** As part of our ongoing household contact study in Kampala, Uganda, we conducted a genome-wide association study (GWAS) of TB severity measured by TBScore, in two independent cohorts of culture-confirmed adult TB cases (n=149 and n=179). We identified 3 SNPs (P<1.0 × 10-7) including one on chromosome 5, rs1848553, that was GWAS significant (meta-analysis p=2.97×10-8). All three SNPs are in introns of RGS7BP and have effect sizes corresponding to clinically meaningful reductions in disease severity. RGS7BP is highly expressed in blood vessels and plays a role in infectious disease pathogenesis. Other genes with suggestive associations defined gene sets involved in platelet homeostasis and transport of organic anions. To explore functional implications of the TB severity-associated variants, we conducted eQTL analyses using expression data from Mtb-stimulated monocyte-derived macrophages. A single variant (rs2976562) associated with monocyte SLA expression (p=0.03) and subsequent analyses indicated that SLA downregulation following MTB stimulation associated with increased TB severity. Src Like Adaptor (SLAP-1), encoded by SLA, is highly expressed in immune cells and negatively regulates T cell receptor signaling, providing a potential mechanistic link to TB severity.

**Conclusions:** These analyses reveal new insights into the genetics of TB severity with regulation of platelet homeostasis and vascular biology being central to consequences for active TB patients. This analysis also reveals genes that regulate inflammation can lead to differences in severity. Our findings provide an important step in improving TB patient outcomes.

## Introduction

Pulmonary tuberculosis (TB) caused more deaths per year than any other pathogen prior to the COVID-19 pandemic [1]. It is the leading cause of death among people infected with human immunodeficiency virus (HIV)[2]. Although incidence is decreasing globally, TB is re-emerging in Sub-Saharan Africa and Southeast Asia[3]. The bacterium, *Mycobacterium tuberculosis* (MTB), causes most TB and is transmitted via airborne droplets from coughing and sneezing by people with active disease. Therefore, it can be a very mobile pathogen in the age of frequent global travel, making global exposure high; between one fourth and one third of the entire global population is latently (asymptomatically) infected. However, far fewer people develop active disease than are infected. In 2020, only 10 million people had active disease with 1.5 million people dying of it [1].

As compared to studies of disease susceptibility or resistance, few studies have focused on TB severity, an important determinant of transmission, morbidity, mortality, as well as disease experience and quality of life [4, 5]. Determinants of TB severity remain uncertain due to heterogeneous definitions. Measures of severity include the TBscore (a validated outcome based on 11 clinically relevant symptoms, Supplemental Table 1), bacillary load, and radiologic findings (e.g. enumeration of pulmonary lesions or area, presence of cavitation) [6-17]. In this study, we chose to use TBscore as it is: 1) based on simple measures of relevant and meaningful clinical parameters that can be ascertained in the resource-limited environments where TB is most prevalent; 2) has previously been validated through comparisons to other measures of TB disease progression and severity [18-20]; and 3) at presentation, it is predictive of mortality in TB patients receiving treatment, associated with quality of life, and even a one point increase is clinically meaningful in some contexts [18-22]. As such, TBscore presents several advantages in terms of ease of measurement and prediction of clinical outcomes. Earlier genetic studies of severity phenotypes focused on candidate genes and not genome-wide analyses, making it likely additional genetic variants elsewhere in the genome associate with TB severity. Notably, some of the severity associating genes (i.e., *IFNG, SLC11A1, MCP1, TLR* variants, and *HLA* variants) are similar or identical to those implicated in previous studies of susceptibility or resistance to TB[23]. However, the overlap in genes for risk and severity may simply be due to the fact that candidate gene studies tended to choose the same genes for the two phenotypes. In contrast, genome-wide analyses may better define how the genetics of TB risk or progression do or do not correlate across severity definitions and may inform distinct functional aspects of TB host genetics.

Understanding the link between DNA variants and RNA expression can inform an understanding of immunological responses and TB pathogenesis. Directly linking genetic variation to immunological function is critical in identifying valid targets for new therapeutics and vaccines[24-28]. This understanding can be partially achieved using bioinformatic databases that annotate function of specific genotypes by connecting tissue specific gene expression or aggregating SNPs from analyses into functional pathways (e.g., GSEA) and can also be achieved by eQTL studies [27, 29-32]. We identified nine studies that have tied gene expression to tuberculosis phenotypes, but only four of them assessed the role of DNA variants in the regulation of gene expression, and none examined the regulation of gene expression in the context of clinical TB severity (Supplemental Table 2). While bioinformatic databases can provide valuable information, they are limited because the cells and tissues available are not always the most relevant to a given phenotype (e.g. TB severity) and they are not under conditions that recapitulate disease related exposures (e.g. active TB) [32]. Further, some eQTL’s are only associated with gene expression in specific contexts, such as an active MTB infection. In the case of MTB infection, resident alveolar macrophages and recruited monocyte-derived macrophages are early targets of MTB in the lung, making these cells informative with respect to the regulation of gene expression in the context of active infection[33]. This situation has, however, yet to be explored[34, 35]. Thus, datasets containing MTB stimulated macrophages can provide functionally relevant information on how variants associated with severity affect RNA expression during MTB infection *in vitro*.

In the present study, we first conducted a case-only genome-wide association study of TBscore to identify variants associated with TB severity. We then followed-up these associated loci in a different set of subjects without active TB disease, utilizing data from monocyte-derived macrophages before and after *in vitro* stimulation with MTB to observe how severity associated variants from our GWAS analysis associated with changes in gene expression in the context of active infection. This study addresses the aforementioned gaps in the current literature on active TB severity by 1) studying the genomic underpinnings of TB severity using a *meaningful, replicable, and validated clinical phenotype*; and 2) *bridging the gap between genetic variants and immunological function* by studying gene expression in the macrophage response, as well as that of other immune cells. Our underlying hypothesis is that genomic variation in humans affects the immunological response to active TB disease, as measured by gene expression, and that this correlates with clinical severity.

## Results

### Study Population

To discover the major genetic variants associated with TB severity, we examined 328 subjects with pulmonary TB (Table 1, 149 subjects in Cohort 1 and 179 in Cohort 2). There were statistically significant differences between the cohorts with respect to HIV status and TBscore, with Cohort 2 having more HIV+ subjects and a lower average TBscore. Both cohorts included more males than females, most subjects were HIV-, and the average age was just under 29 in both cohorts.

**Table 1.**
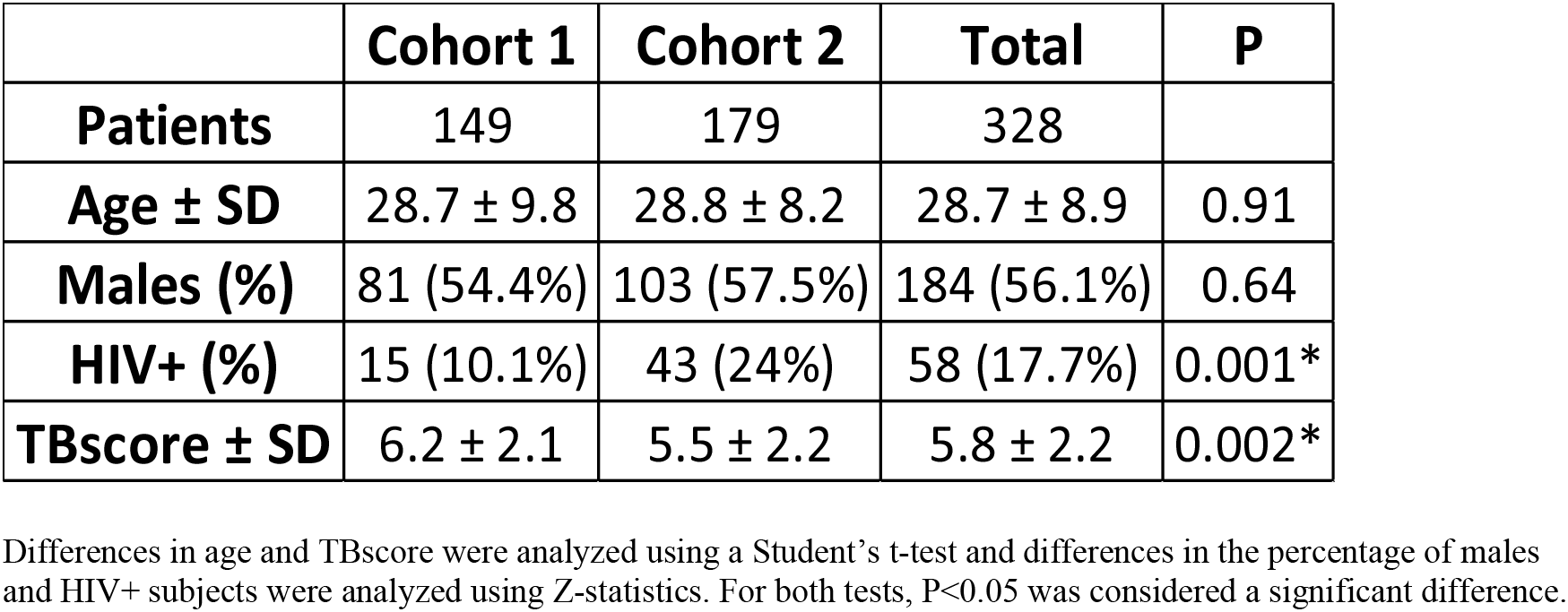
Cohort Characteristics.

|  | **Cohort 1** | **Cohort 2** | **Total** | **P** |
| --- | --- | --- | --- | --- |
| **Patients** | 149 | 179 | 328 |  |
| **Age ± SD** | 28.7 ± 9.8 | 28.8 ± 8.2 | 28.7 ± 8.9 | 0.91 |
| **Males (%)** | 81 (54.4%) | 103 (57.5%) | 184 (56.1%) | 0.64 |
| **HIV+ (%)** | 15 (10.1%) | 43 (24%) | 58 (17.7%) | 0.001* |
| **TBscore ± SD** | 6.2 ± 2.1 | 5.5 ± 2.2 | 5.8 ± 2.2 | 0.002* |

Differences in age and TBscore were analyzed using a Student's t-test and differences in the percentage of males and HIV+ subjects were analyzed using Z-statistics. For both tests, $P < 0.05$ was considered a significant difference.

### Genome-Wide Association Results

Individually, the two cohorts showed no sign of genome-wide inflation (Supplemental Figures 5, 6) in their Q-Q plots or genomic control statistics (λ<1.0). PCA analysis revealed that none of the top 10 PC’s were associated with TBscore in either cohort. Based on this and the low amount of variation explained by the PC’s, we decided not to include them in the initial regression equation (Supplemental Figures 2, 3). The Q-Q plot for the full range of meta-analytic P-values appears to deviate from the line but the genomic control parameter (λ) was 0.98, indicating little to no genome-wide inflation (Figure 1). Neither cohort showed any SNPs that were GWAS significant when considered individually (Supplemental Figures 7, 8). There were a total of 11,323 SNPs showing an association with P<0.05 in both cohorts and beta values with the same direction of effect (i.e. both negative or both positive) (Supplemental Tables 3 and 4). Of these, 10,750 SNPs passed the I^2^ threshold for heterogeneity. Out of the 10,750, there was one SNP on chromosome 5 (rs1848553) that was GWAS significant with a meta-analytic P-value of 2.97×10^−8^ and a beta of -0.97 (Table 2, Figure 2). Two other SNPs in this gene are close to genome wide significant as well (p = 7.78×10^−8^ for both, Table 2). We conducted sensitivity analysis to evaluate the impact of PCs, and this analysis demonstrated that the association of rs1848553 at a GWAS significant level is not sensitive to the PC’s being in the regression equation in the analyses (Supplemental Figures 2,3, Supplemental Methods).

**Table 2.** Annotation for Top 10 SNPs for Association with TBscore, Sorted by P-Value.

| **SNP** | **CHR:BP** | **Ref/Alt** | **Gene** | **Location** | **Meta-analytic P-Value** | **Meta-analytic $\beta$** | **MAF in Cohort 1** | **MAF in Cohort 2** |
| --- | --- | --- | --- | --- | --- | --- | --- | --- |
| rs1848553 | 5:63805731 | C/T | *RGS7BP* | Intron | $2.97 \times 10^{-8}$ | -0.97 | 23.7% | 25.1% |
| rs6870654 | 5:63831964 | T/C | *RGS7BP* | Intron | $7.78 \times 10^{-8}$ | -0.83 | 41.7% | 37.1% |
| rs6894580 | 5:63832264 | G/A | *RGS7BP* | Intron | $7.78 \times 10^{-8}$ | -0.83 | 41.7% | 37.1% |
| rs2829189 | 21:25966515 | A/T | None | CTCF Binding Site | $1.30 \times 10^{-7}$ | 1.23 | 12.5% | 13.3% |
| rs60496505 | 5:63840208 | A/C | *RGS7BP* | Intron | $1.83 \times 10^{-7}$ | -0.80 | 43.0% | 38.3% |
| rs11210569 | 1:38805929 | T/C | *LOC105378657* | Intron | $1.89 \times 10^{-7}$ | 0.89 | 26.3% | 26.2% |
| rs6873254 | 5:63823469 | A/G | *RGS7BP* | Intron | $1.98 \times 10^{-7}$ | -0.79 | 37.5% | 37.9% |
| rs4816976 | 21:25967403 | C/T | None | Intergenic | $2.04 \times 10^{-7}$ | 1.19 | 12.8% | 13.3% |
| rs2829200 | 21:25978459 | G/C | None | Intergenic | $2.58 \times 10^{-7}$ | 1.23 | 10.2% | 13.0% |
| rs11208579 | 1:65534491 | A/G | *JAK1* | Promoter | $2.91 \times 10^{-7}$ | 1.33 | 8.6% | 13.8% |

Location and minor allele frequency in the 1000G project were ascertained from Ensembl Genome Browser v104. Ref/Alt refer to the reference and alternative alleles (which were the major and minor alleles, respectively). Copies of the alternative allele are what the beta value is showing in the regression equation.

**Figure 1.**
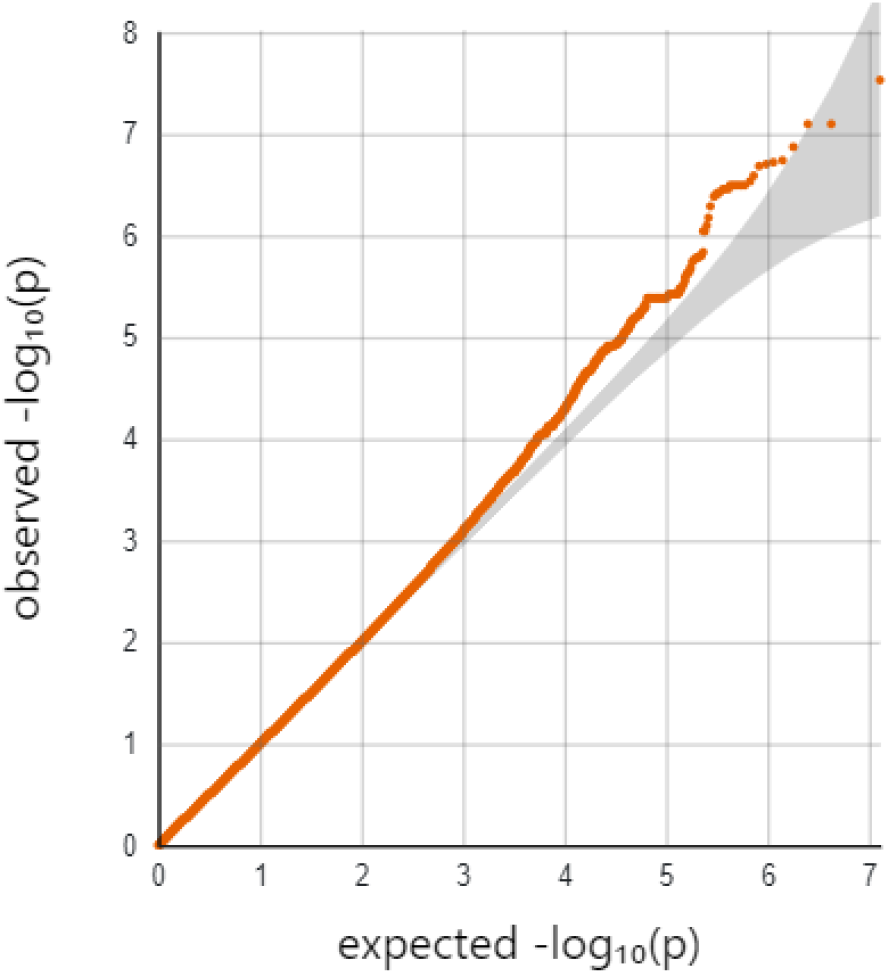
Quantile-Quantile Plot for Meta-Analytic P-values of Association Between SNPs and TBscore. The quantile-quantile (Q-Q) plot shows the inverse log(10) of the observed p-values on the Y-axis relative to what is expected if there was no association on the x-axis. Deviations above the line indicate an association with the outcome. If the line deviates at the low quantiles, then this is considered evidence to suggest genome-wide inflation of the test statistics, which typically indicates unmeasured confounding (λ=0.98).

**Figure 2.**
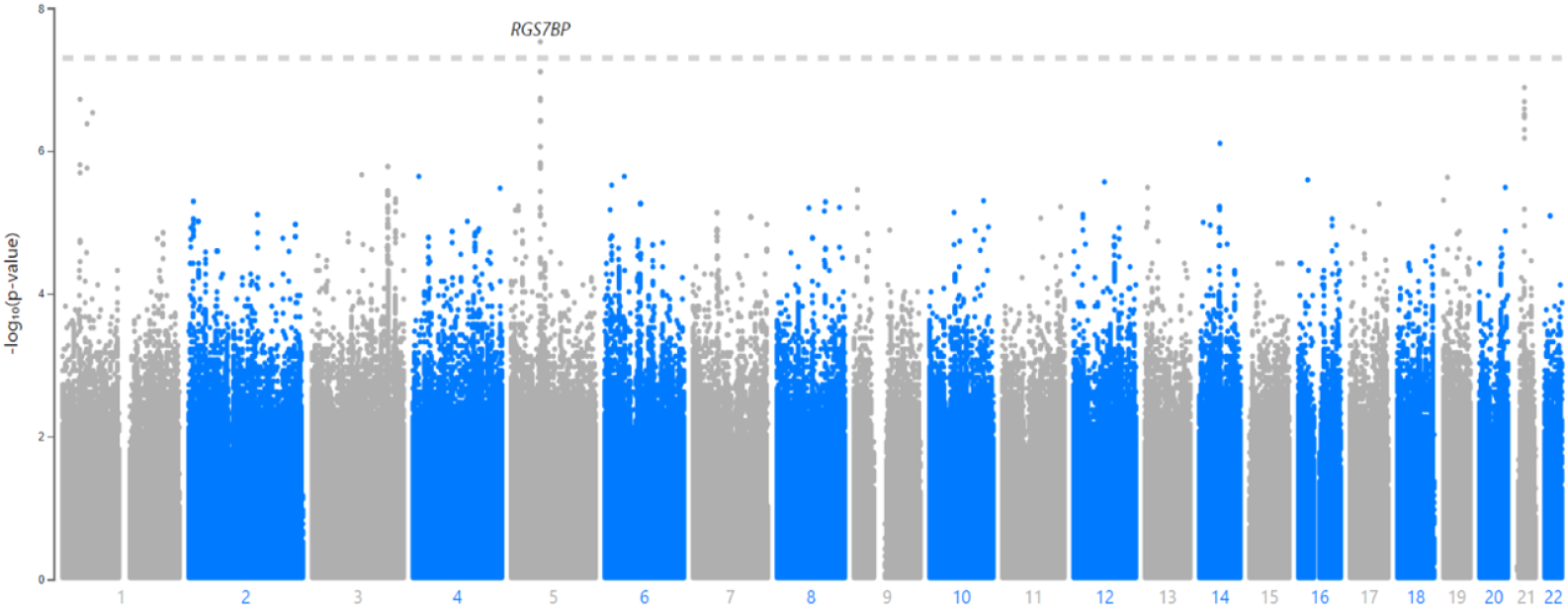
Manhattan Plot for Meta-Analytic P-Values of Association Between SNPs and TBscore. The Manhattan plot shows the inverse log(10) of the p-values for the association between each SNP and TBscore on the y-axis and the x-axis represent the physical location of each SNP on the chromosomes, which are in order from 1-22.

### Annotation for GWAS Significant SNP: rs1848553

SNP rs1848553 is a C to T allele change, the MAF in the two cohorts combined was 24.7%, and did not vary much between cohorts (Supplemental Table 5). The T allele is generally very rare in 1000G reference populations. However, in African populations, the T allele is much more common (21%) than in other continental populations, where the T allele ranged from 0-2.6% (Supplemental Table 6). The beta value indicates each copy of the minor allele is associated with just under a 1-point decrease in the TBscore in the additive model. This represents a clinically meaningful reduction in severity [18], as well as an important reduction in the risk of mortality. Considering each T allele is associated with nearly a 1-point reduction in TBscore, T/T homozygotes have nearly a 2 point reduction in TBscore relative to C/C homozygotes (Figure 4). The beta and P-values did not vary across cohorts and the I^2^ for this SNP was <1%, indicating little to no heterogeneity. The GWAS significant SNP, rs1848553, is located within an intron of the protein-coding *RGS7BP* gene [36]. There are 18 other SNPs in the same area (63.6Mb to 64.0Mb) on chromosome 5 that show an association with TBscore with P<1×10^−5^, including the 2 SNPs that had a P<1×10^−7^ (Figure 3; Table 2). *RGS7BP* codes for a protein that regulates trafficking of G-proteins between the nucleus and the plasma membrane [37].

**Figure 3.**
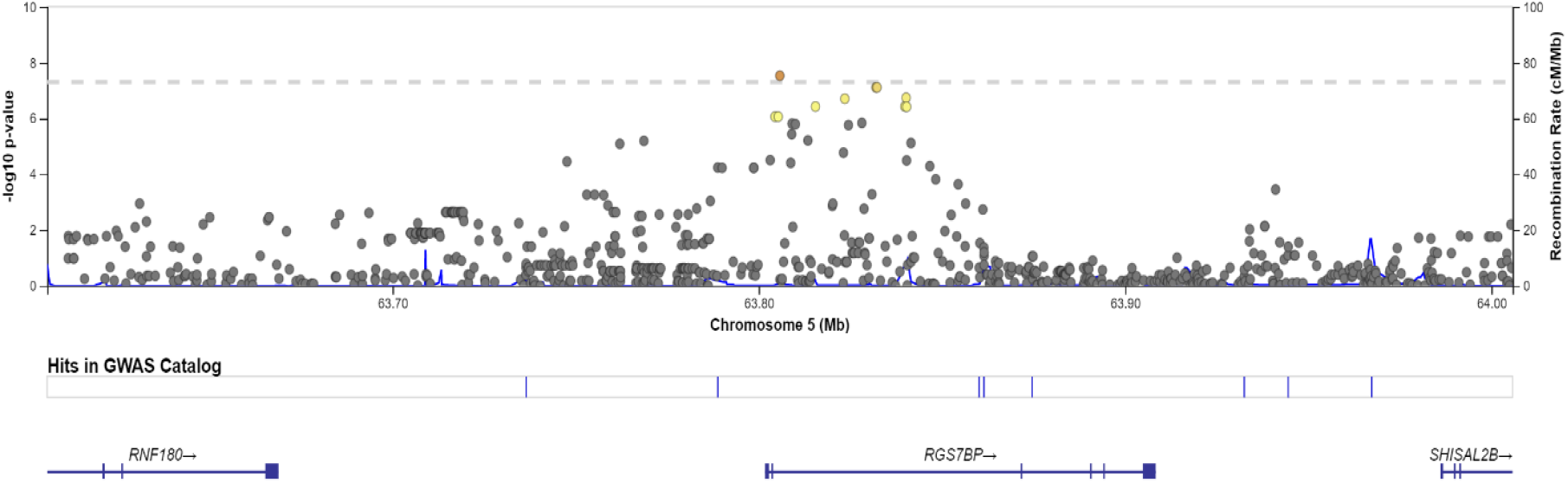
LocusZoom Plot for Region Surrounding rs1848553. The LocusZoom plot shows the region surrounding rs1848553 on chromosome 5, using an LD panel and reference genome from the AFR super-population in the 1000G project. Yellow and orange indicate higher levels of LD.

**Figure 4.**
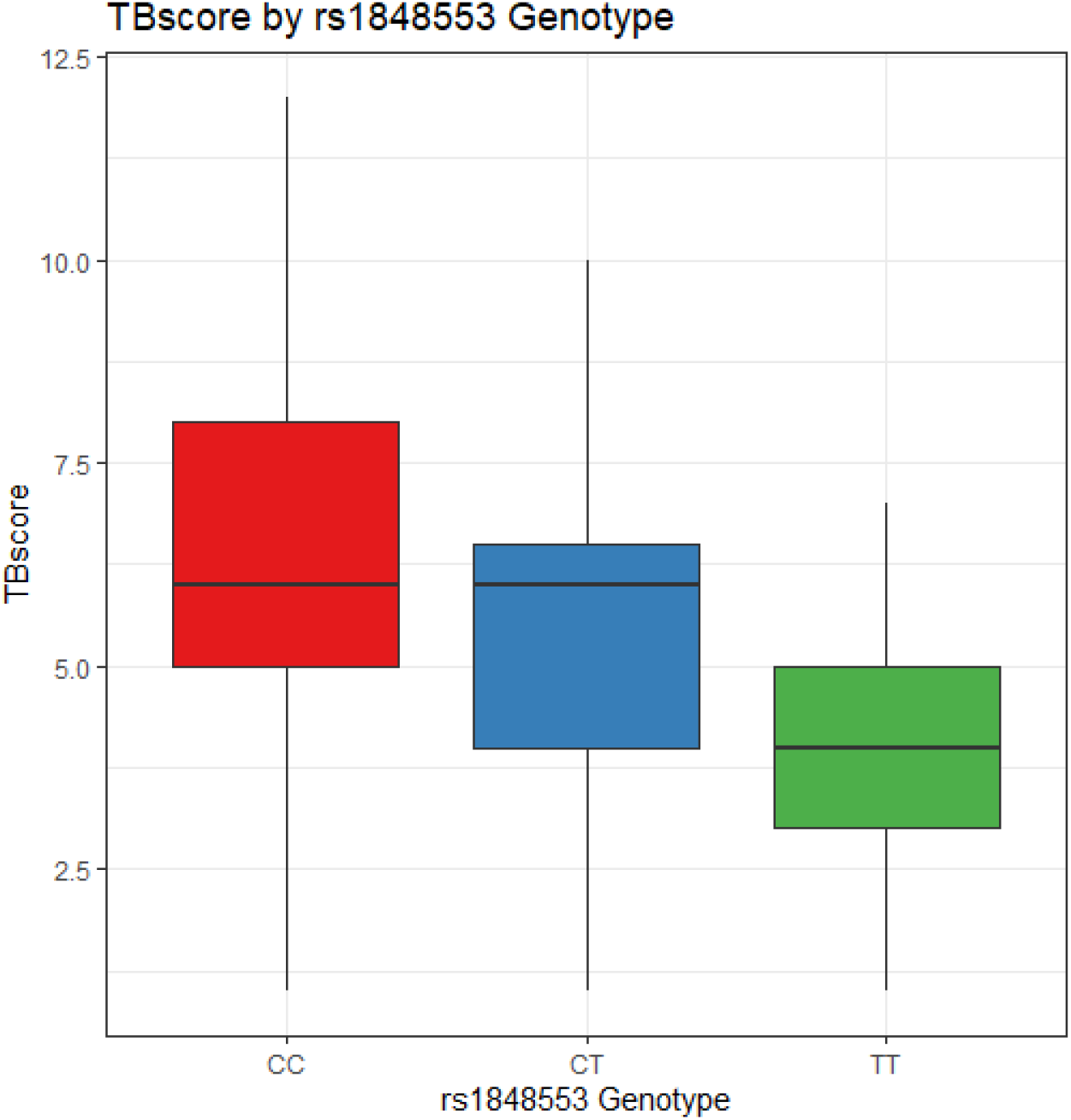
Boxplot of TBscore by rs1848553 (GWAS significant SNP) Genotype. Distribution of TBscore by genotype

### Chest X-Ray Analysis

Of the 169 SNPs showing an association with TBscore at P<1 ×10^−5^, 165 of these showed the same direction of effect in the CXR analyses: i.e., the beta value for association with TBscore was <0 and the OR for association with CXR extent was <1, or the TBscore association beta was >0 and the CXR OR was >1 (Supplemental Figures 9 and 10, Supplemental Table 3). While the direction of effect was consistent in 97.6% of the SNPs that showed an association with TBscore at the P<1 ×10^−5^ threshold, 20 of those 165 SNPs showed an association with CXR extent with P<1 ×10^−5^. Thus, the consistency in the direction of effects provides evidence that the TBscore is a similar and valid measure for quantifying clinical severity.

### Decile Regression Analyses

The decile regression for rs1848553 and TBscore was consistent with a negative or zero beta value for the whole range of the TBscore (Supplemental Figure 11). This analysis shows that for effect of rs1848553 genotype, there is a stronger effect at the highest end of the TBscore deciles than anywhere else but the effect never reverses direction. Thus, it appears that genetics has the greatest influence on severity among the subset of active TB patients who have most severe or most mild disease, and it makes the least impact on those with TBscores close to the mean.

### Examination of Prior Published Results Related to Susceptibility and Severity

We examined our genome-wide TBscore association results for 12 previously identified TB susceptibility SNPs and 3 SNPs in our data showed association with TBscore gene regions associated with disease (P<0.05) (Supplemental Table 7)[38]. *IFNG, TLR4*, and *VDR* all showed P<0.05 for the association with TBscore, and *SLC11A1* had P=0.053. Most of the gene regions examined did not show a statistically significant association, indicating that there are likely unique sets of genes that drive severity versus susceptibility. To follow-up our previously identified association with *IL12B* variants [17], we examined the association between any SNPs +/- 50kb from *IL12B* with P<0.05 for association with TBscore in Cohort 1, and identified three SNPs with P<0.05 in this region (Supplemental Table 8). These findings add further evidence that there are likely distinct sets of genes associated with susceptibility to and severity of TB disease.

### Enrichment Analyses

A total of 169 SNP associations had P<1 ×10^−5^ and were used for annotation and enrichment analysis [39-41]. The analyses showed enrichment for two Reactome pathways: platelet homeostasis and organic anion transport (Table 5). The MAGMA analysis of gene-level associations with TBscore did not show any significant single gene results at the genome-wide level; the most significant association (P=2.1×10^−3^) was in the *PSORS1C2* gene, also known as the psoriasis susceptibility 1 candidate 2 gene that is thought to confer susceptibility to psoriasis. There were 19,220 genes represented in the summary statistics and the resulting p-value threshold for genome-wide significance at the gene-level was P<2.6×10^−6^. While no individual genes or gene sets were found to be significant, the MAGMA gene level analysis of tissue specificity showed that the genes represented in the data are significantly differentially expressed, and specifically up-regulated, in blood vessels based on tissue specificity data from GTEx v8 (Figure 5).

**Table 5.**
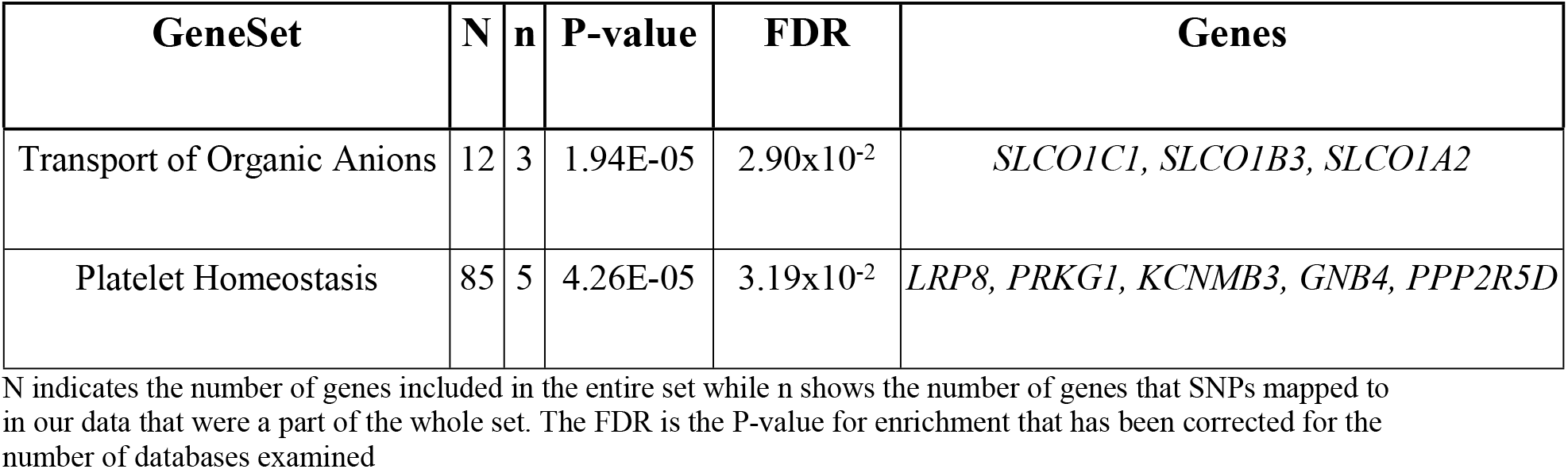
Reactome Gene Set Enrichment Results for SNPs P<1×10^−5^.

**Figure 5.**
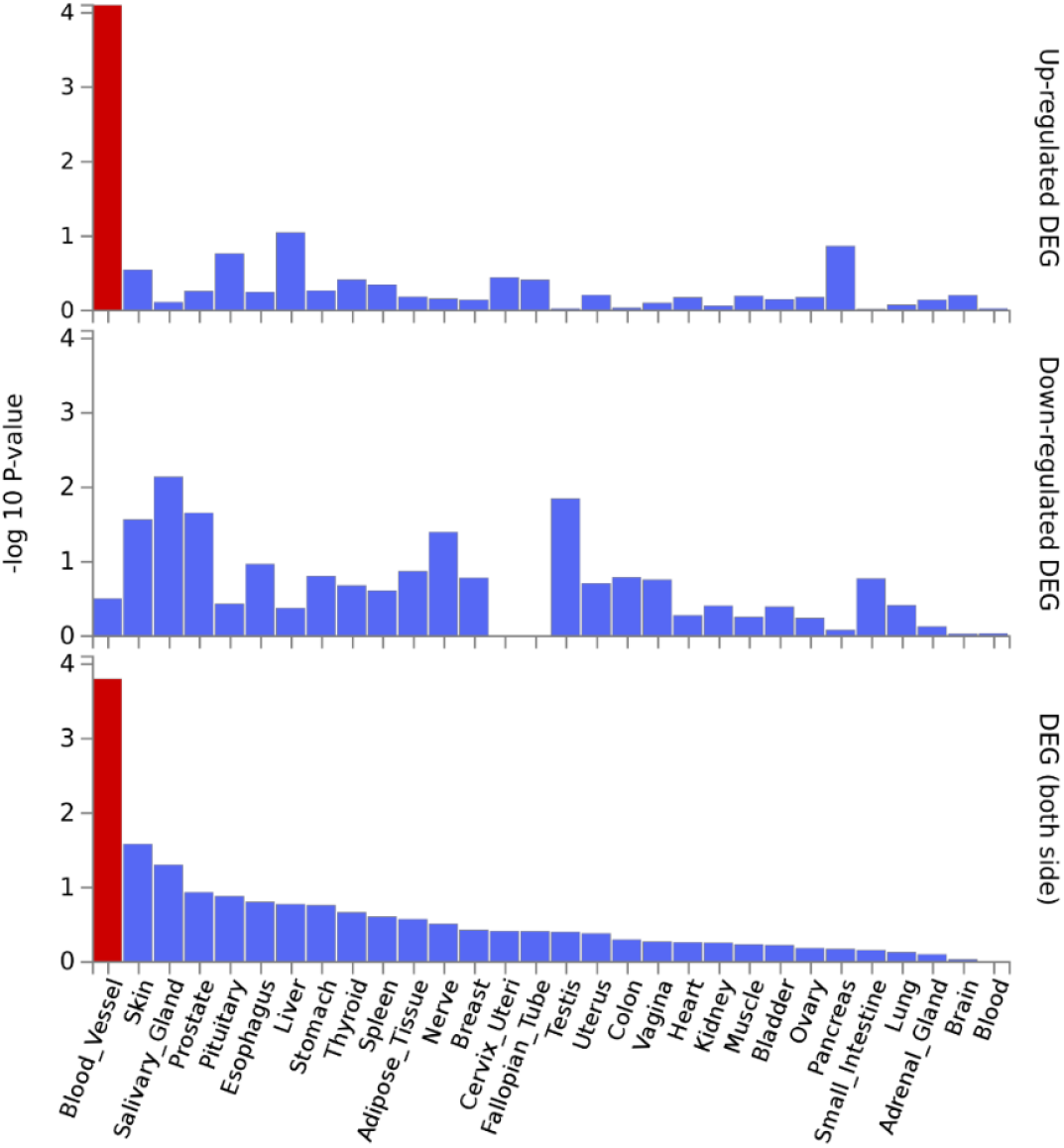
Gene-level tissue Specificity from MAGMA Analysis and GTEx Database. FUMA GWAS uses MAGMA gene-level analyses and differential gene expression data from GTEx v8 to determine if the genes to which the SNPs are mapped are significantly differentially expressed in any tissues. This analysis showed that the mapped genes were significantly upregulated in blood vessels (indicated by the red color).

Using FUMA to determine if any of the 169 SNPs from our GWAS analysis were eQTL’s, we found that 25 of the SNPs were eQTL’s that associate with expression of 28 genes; some of the eQTL’s showed an association with more than one gene. There were a total of 150 eQTL effects across different tissues and databases, as many of the 25 eQTL’s associated with expression in multiple tissues and/or databases (Supplemental Figure 12 shows the 28 genes). Interestingly, 5 of the SNPs with P<1×10^−5^ were cis eQTL’s for *RGS7BP* in the same region on chromosome 5 as the top GWAS hit. This is consistent with the annotations that showed it is a known regulatory variant. The STRING database analysis of protein-protein interactions (PPIs) did not show significant enrichment for interactions or for any pathways (Supplemental Figure 12).

### eQTL Analysis of Severity Associated Variants

We next examined whether these suggestive TB severity SNPs were eQTLs in Mtb-stimulated monocytes (cohort characteristics in Supplemental Table 9). There were four SNPs that showed significant SNP by stimulation interactions that were significant with FDR corrected P<0.1, and all of these were cis-eQTL’s (Table 6, Supplemental Table 11). Upon examining the marginal effects (no interaction term in model) for these four significant SNPs, only rs2976562 at the Src-like adaptor *(SLA)* gene was significantly associated with expression in the MTB-stimulated stratum (Table 7). This eQTL did not have a significant effect in media-only. Thus, rs2976562 meets the definition of a stimulation dependent eQTL, as it is active only within the context of MTB infection. In the analyses that were stratified by RSTR or LTBI status, this SNP showed a similar Beta (−0.71), but were not statistically significant (FDR=0.15), likely due to the decreased sample size. This indicates that the relationship between rs2976562 and *SLA* is similar between RSTR and LTBI, and thus, robust to differences in patients’ clinical characteristics, despite only being significant when the data are combined. Notably, rs2976562 showed interaction with MTB stimulation that associated with both *SLA* and *NDRG1* expression, but the association with *NDRG1* was not significant after FDR correction (Table 6). However, rs2976562 was also found to be an eQTL for *NDRG1* in the eQTLGen cis eQTL’s database during our interrogation of publicly available eQTL databases described above (Supplemental Figure 12, Supplemental Table 10).

**Table 6.** Severity Associated cis-eQTL’s with P<0.05 for Effect of Interaction Between Stimulation and Genotype on Expression.

| SNP | Gene | P | FDR | Beta |
| --- | --- | --- | --- | --- |
| rs58648494 | *ANKRD33B* | 0.004 | 0.063 | 0.95 |
| rs8100115 | *ICAM1* | 0.0041 | 0.065 | 0.81 |
| rs8100115 | *DNMT1* | 0.0044 | 0.065 | -0.44 |
| *rs2976562 | *SLA* | 0.0049 | 0.068 | -0.67 |
| rs8100115 | *P2RY11* | 0.025 | 0.27 | -0.79 |
| rs2976562 | *NDRG1* | 0.029 | 0.28 | 0.63 |
| rs4459669 | *KLF16* | 0.032 | 0.28 | -0.25 |
| rs11210569 | *MTF1* | 0.037 | 0.29 | 0.38 |

The p-value and beta is for the interaction between SNP and MTB stimulation and the gene column shows the gene for which the interaction is associated with expression
\*Indicates eQTL that was also significantly associated (P<0.05 after FDR correction) with expression in MTB-stimulated cells (Figure 6)

MTB stimulation increases the expression of *SLA* for individuals with CC and CT genotypes (Figure 6). The T allele for rs2976562 is associated with lower expression of *SLA* after stimulation with MTB, but is associated with a relative increase in *SLA* expression prior to MTB stimulation (p=0.03). Hence, the effect of rs2976562 is dependent on MTB infection status of the cells. Comparing this to the boxplot of the relationship between rs2976562 and severity shows that this same allele is associated with increased severity, especially among homozygotes, who appear to have very severe disease on average (Figure 7). Thus, there appears to be a relationship between downregulation of *SLA* upon MTB stimulation and more severe TB disease among T/T homozygotes.

**Figure 6.**
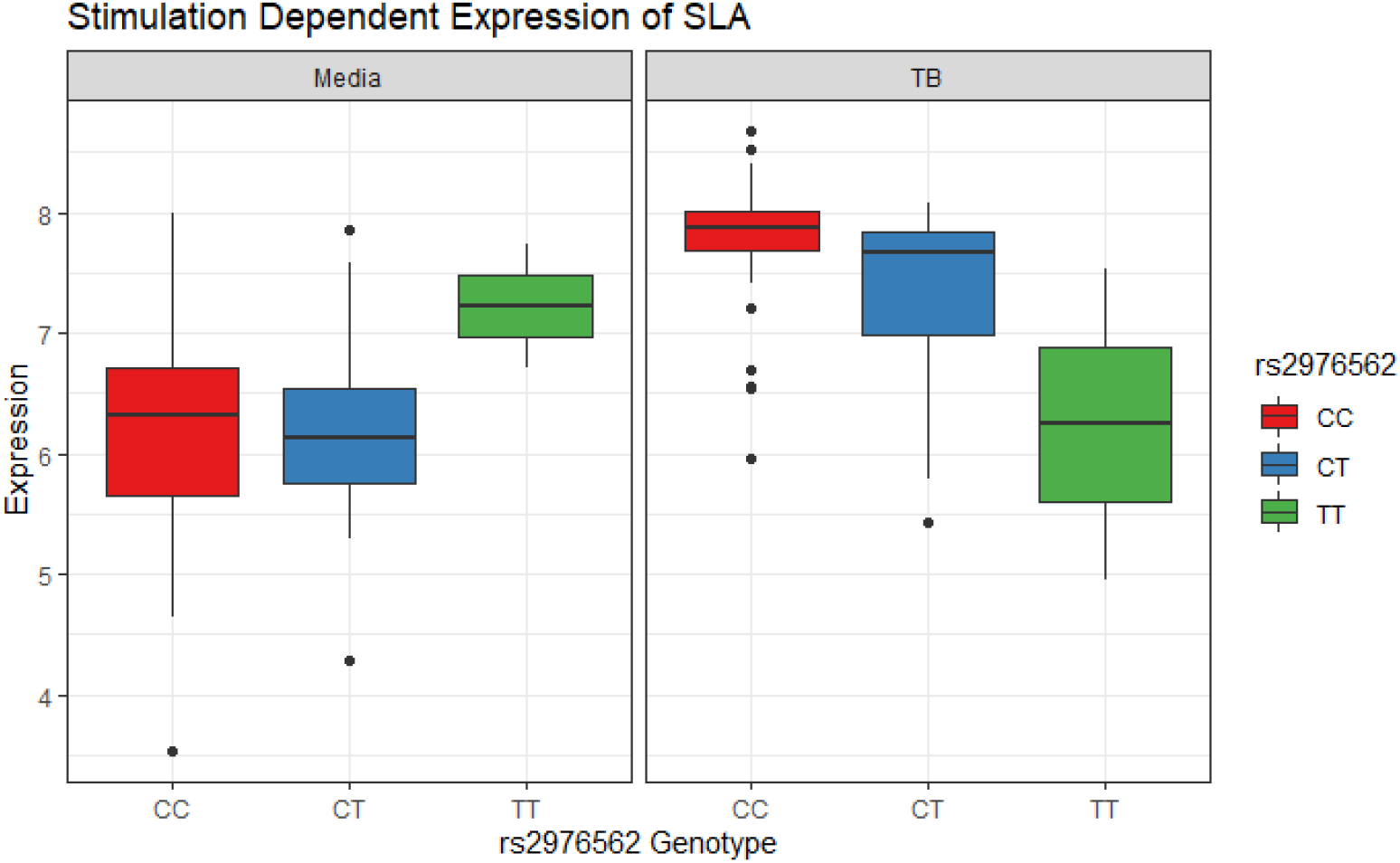
Relationship between rs2976562 alleles and SLA Expression Before and After Stimulation with MTB. rs2976562 Genotype Distribution: 96 subjects CC, 44 CT, and 4 TT. P value in media=0.41, β in media=0.6, P-value in MTB stimulated=0.03, β in MTB stimulated=-0.32

**Figure 7.**
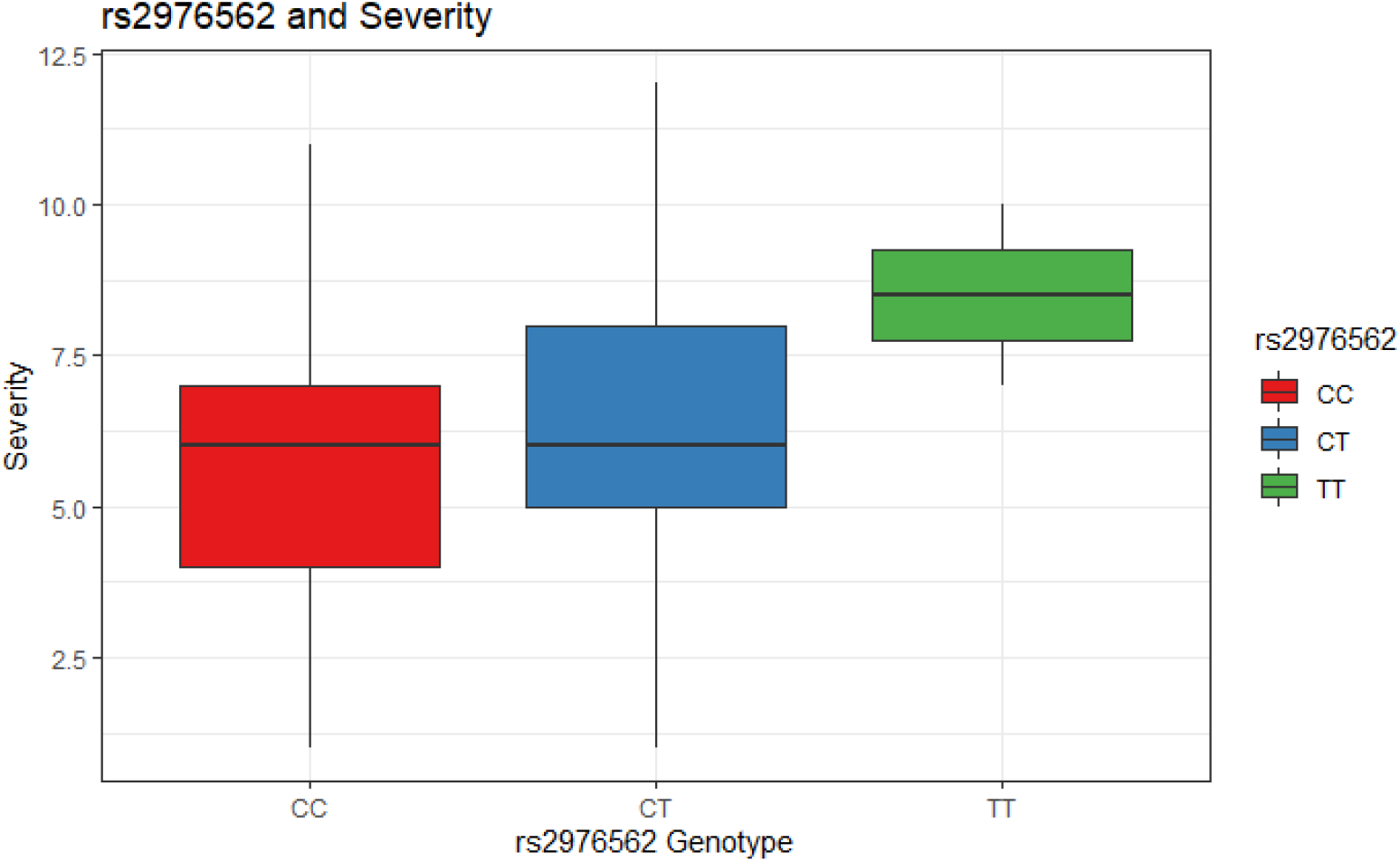
Relationship between rs2976562 alleles and TBscore (severity) Distribution of TBscore by genotype

The data show that a statistically significant relationship between *SLA* expression and rs2976562 genotype only becomes apparent in the context of MTB stimulation, though the T/T homozygotes do appear to have higher expression of *SLA* prior to stimulation but the relationship was not statistically significant. An examination of population genetic data shows that the T allele is found at lowest frequency (11%) in African populations, but it is actually the major allele in Asian and American populations (60-70%) and common in European populations [42] (Supplemental Table 12).

## Discussion

Overall, our results show that human genetic variation is associated with TB severity. We found a single SNP that was GWAS significant, but enrichment analyses and tissue specificity both revealed variation in several processes related to vascular biology that have been previously implicated in the inflammatory response to infectious disease. Our eQTL analysis results suggested that human genetic variation in an important aspect of antigen presentation may affect the host response to infection with MTB. A better understanding of these mechanisms, and how they relate to severity and mortality in active TB patients, may lead to greater therapeutic insight that can reduce the negative impact of active TB disease on human health.

While the MAGMA analysis did not show any statistically significant single-gene effects, the tissue expression analysis of our MAGMA results showed that, collectively, the genes represented in our severity-associated results are significantly up-regulated in blood vessels in response to MTB infection. The vascular endothelium plays an important role in thrombosis and inflammation, and the vasculature is responsible for enabling the extravasation of immune effector cells in the response to infection[43, 44]. Acute changes in blood pressure during active infection can lead to organ failure and death in COVID-19 (hypertension) and acute sepsis (hypotension) [45, 46]. Further, vasculitis and stroke (particularly in the context of TB meningitis) have been posited as complications of TB disease as a result of inflammation and dysregulation of vascular function[47-49]. If differences in inflammation, coagulation, and regulation of vascular function lead to more severe outcomes and/or death, then an understanding of this phenomenon, and how to address it, could drive an improvement in outcomes and reduction in mortality for active TB patients.

Previous studies of TB susceptibility identified genes that are primarily involved in the host immune response. However, the genetics of severity appears to incorporate a different biological process, namely platelet homeostasis. Specifically, platelet homeostasis was found to be enriched in the GSEA analyses using FUMA, and platelets are an important part of the response to both inflammation and infection[50, 51]. Platelets are involved in crosstalk between immune effector cells and aid in the body’s ability to sense pathogens and enable infection-induced inflammation[50-53]. This inflammation often leads to a state that boosts coagulation in humans and a previous study showed that TB patients are in a pro-coagulatory state[54, 55]. The damage that dysregulation of platelet homeostasis and coagulation can cause has also been demonstrated in acute sepsis and septic shock [56]. Notably, septic shock has been‘ previously reported as a common cause of death in pulmonary TB patients[57].That most genes previously associated with susceptibility did not significantly associate with TB severity emphasizes that although the two phenotypes have some overlap thet are likely genetically and biologically distinct.

Our eQTL results help clarify the effect that some of the severity associated DNA variants exert on gene expression. While the GWAS significant SNP was not shown to be an eQTL, multiple SNPs within the same region as the GWAS significant SNP (rs1848553) appear to be eQTL’s for *RGS7BP*, implying that there may be a functional role for this gene in active TB. The stimulation-dependent eQTL, rs2976562, was an important regulator of *SLA* in the context of *in vitro* stimulation in monocytes and resides in a flanking promoter region[58]. While this region does not contain the promoter for the *SLA* gene, it has been associated with expression of other genes and may be an enhancer for *SLA*.

The *SLA* (Src-like adaptor) gene codes for the SLAP-1 protein. The DICE database indicates that *SLA* is expressed in a number of immune cells, including monocytes, but that its expression is highest in T-cells (Supplemental Figure 13)[59]. SLAP-1 is an adapter protein that negatively regulates T-cell receptor signaling, inhibits T-cell antigen-receptor induced activation of nuclear factor of activated T-cells, and is involved in the negative regulation of positive selection and mitosis of T-cells[37, 60, 61]. SLAP has a role in activation and maturation of monocyte and dendritic cells through downregulation of granulocyte macrophage colony-stimulating factor receptor (GM-CSFR) [35]. SLAP deficient bone marrow-derived dendritic cells produce less TNF-α and IL12 in response to LPS, fail to stimulate T-cells in mixed lymphocyte reactions, and are less effective at inducing IFN-γ secretion from T cells[62]. Thus, a deficiency of SLAP-1 likely impairs a robust immune response, and reduces ability to generate cytokines (IL12, IFN-γ, and TNF-α) that are important drivers of the host immune response to active TB infection[63-65]. Further, variants that code for these proteins have previously been implicated in TB severity and/or susceptibility. Our results in conjunction with prior literature on the role of *SLA* indicate that decreased *SLA* expression may be associated with more severe disease and this may be explained by dysregulation in the maturation of monocytes and dendritic cells.

This study is not without limitations: 1) our sample size is small by the standards of many modern GWAS studies; and 2) there were differences between the two cohorts, namely in TBscore and in the proportion of HIV+ individuals. The analyses controlled for the latter factor by adjusting for HIV status. That said, the genomic control parameter was below 1, so it does not appear that there was significant genome-wide inflation. Furthermore, we used a stringent I^2^ threshold to exclude SNPs showing heterogeneity of effects. Thus, it is unlikely that differences between the two cohorts substantially affected the associations. For our eQTL analyses, a more directly related phenotype would have been cells harvested from patients with active TB, as this was the patient population from our GWAS analysis. However, the stratified analysis indicated that the association we observed was similar in LTBI patients, and thus unlikely to be sensitive to the inclusion of RSTR subjects in the data. The small sample size also affected our eQTL analysis; the strongest association between rs2976562 genotype and *SLA* expression was among TT homozygotes, of which there were only 4.

In conclusion, this study demonstrates that TB severity associates with genetic variation and indicates that variation in both the regulation of platelet homeostasis and vascular function may be driving outcomes for active TB patients. Our eQTL results indicated that regulation of *SLA* may be an important driver of variation in active TB severity due to an impact on a fundamental aspect of the host immune response to TB disease. Further study of the impact of *SLA* may yield insight into a more effective host immune response that is associated with less severity and mortality. Future studies should consider the role that infection-induced inflammation plays in active TB severity, in the hope that mortality and other severe outcomes might be mitigated or avoided through a better understanding of these processes.

## Methods

### Ethics statement

The study protocol was approved by the National HIV/AIDS Research Committee and the institutional review board at University Hospitals Cleveland Medical Center. Final clearance was given by the Uganda National Council for Science and Technology. All participants provided written informed consent.

### Study participants

TB cases and extensive clinical data were ascertained as part of the Kawempe Community Health Study (KC Study) [66], a longitudinal household contact study conducted in Kampala, Uganda from 2002-2017 through the Uganda-CWRU Research Collaboration. The KC Study enrolled 3,818 total participants, of which there were 872 adult active TB index cases. In the present case-only analysis of severity, a subset, for whom available genotype data and the information necessary for assigning the TBscore exists, were identified from these 872 index cases (Supplemental Table 1). The TBscore developed for adults may not appropriate for individuals under 15. Thus, our sample was limited to TB cases 15 years old and older. We examined two subsets, as described in our past publications, that will be referred to as Cohort 1 and Cohort 2 (N=149 and N=179, respectively) according to different genotyping platforms available at different times during the study (described below). The Bandim TBscore was constructed as described previously [19, 67]; further details are provided in the Supplemental Methods.

All TB cases were culture-confirmed based on isolation of MTB from sputum and clinical characteristics were assessed during the visit at which subjects were diagnosed with active TB. X-rays were performed at the Uganda Cancer Institute. Additional details about the original study protocol are described elsewhere [66]. The two subsets differed in percentage of HIV positive individuals (Table 1); therefore, HIV status was used as a covariate in all regression models. Previous analyses of microsatellite data from these cohorts indicated no substantial population substructure, as confirmed by previous principal components (PC) analyses [17, 68].

### Monocyte Transcriptional Profiles

A follow-up cohort including 72 Ugandan subjects without active TB infection was included in a transcriptome-wide study to assess the association between gene expression and variants we identified in our GWAS. These subjects were ascertained from the KC Study but included only subjects without active TB because this cohort had both genotype and RNA expression data available. The follow-up cohort, without active TB infection, was part of an analysis that examined whole blood and monocytes isolated from highly MTB-exposed, HIV-negative donors in Uganda[69, 70]. Some of these subjects (n=34) showed resistance (classified using TST and IGRA) to infection by MTB after repeated exposure, and are referred to as resisters (RSTR), as previously described [71, 72]. In addition to the RSTR subjects, there were donors with latent tuberculosis (LTBI) defined by concordant positive TST/IGRA. For this study, monocytes were isolated from PBMCs and then stimulated with MTB for 6 hours. RNA expression levels were assessed in these samples using RNA-seq with and without stimulation by MTB[73].

### Genotyping and QC

Cohort 1 was genotyped on the Illumina Infinium Mega^EX^ chip, comprising 2.1M markers genome-wide. For Cohort 2, we used the Illumina HumanOmni5 microarray comprising 4.3M markers genome-wide, offering high genome wide coverage of common genetic variation within African populations[74]. Prior to imputation, only SNPs that had a call rate greater than 0.98, minor allele frequency (MAF) > 0.05, and did not show deviation from Hardy-Weinberg equilibrium (p<10^−6^) before and after imputation in both samples were used in the analysis. The total number of SNPs that overlapped between the two cohorts after imputation quality control (see Supplemental Methods) was 6,421,278. Principal components were computed using Plink v1.9 (Supplemental Figures 2,3).

Our follow-up cohort without active TB used for the analysis of monocyte derived macrophages was genotyped separately from TB cases in Cohorts 1 and 2. The genotyping for this cohort was done on the Illumina Mega^Ex^ Chip and had 1,042,921 SNPs prior to imputation. To maximize the overlap between the list of SNPs associated with TBscore and those included in the genotype data for the 72 subjects with RNA-seq data, we imputed SNP calls on the follow-up cohort with the Michigan Imputation Server (Minimac4) with a Minimac R^2^ >0.3 on the whole MegaEx chip[75]. The 1000G Phase 3 v5 AFR panel was used as the reference population and Eagle2 was used for phasing[76]. After imputation, we restricted the analysis to SNPs with MAF > 0.05, genotyping rate > 98%, and those that did not violate Hardy-Weinberg equilibrium (at p<10^−6^). Gene expression was measured using RNA-seq in monocyte-derived macrophages, as previously described (Supplemental Methods)[70, 73].

### Genome-wide association analysis

To assess the association between genetic variants and TBscore, we utilized a linear regression model with sex and HIV status as covariates in Plink v1.9 software. We then combined the summary statistics from the two cohorts to generate meta-analysis derived p-values. To determine meta-analysis p-values and beta coefficients across the two cohorts, we utilized random effects meta-analysis with inverse variance weighting. Based on the Cochrane handbook recommendations, all variants with an I^2^ > 40% were excluded from the analysis to reduce heterogeneity between the cohorts [77]. To be considered GWAS significant, the association between a variant and TBscore had to have a p<0.05 in both cohorts, the sign of the beta value had to be the same in both cohorts, and the meta-analysis needed to meet the canonical GWAS threshold (*P* <10^−8^). A power calculation showed that, with our sample size and mean TBscore, we had 48% power to detect a difference of 1 point on the TBscore at p=5×10^−8^ and at a minor allele frequency of 0.25, using an additive model. To be included in further enrichment and annotation analyses, the meta-analytic P-value had to be below 1×10^−5^. We chose this latter threshold because previous studies have shown that some variants that do not meet the GWAS threshold may still have important regulatory or biological functions, and may be worthy of further study and follow-up, especially in the context of gene regulation [39-41]. We used FUMA GWAS to annotate and enrich our SNPs below this threshold. Analyses performed with FUMA included gene mapping, regulatory annotation, tissue specificity, MAGMA analysis (gene-based analysis), gene set enrichment, and pathway analyses (Supplemental Methods) [78]. In addition to FUMA, we utilized GeneCards, Ensembl, DICE, and STRING DB to annotate and enrich our results with respect to function, expression, and downstream protein interactions [36, 37, 59, 79].

To look more closely at SNPs that showed GWAS significance, we performed decile regression on all significant SNPs to determine if and how their relationship with TBscore varied across the distribution. Decile regression, performed using QuantReg Software in R 3.6.3, shows how the beta values for the SNPs in the linear regression differ by deciles of the TBscore. The decile regression utilized the same covariates in the regression equation as the GWAS analysis (HIV status and sex).

To validate our GWAS summary statistics showing association with TBscore, we also examined the association between the genetic variants of interest and radiological severity, as determined by extent of disease using chest X-ray (CXR) data. Extent of lung involvement was measured using the US National Tuberculosis and Respiratory Disease Association (US NTRDA) grading system from radiographs taken at the Uganda Cancer Institute and performed on the same patients as the analysis of TBscore (i.e. they were identical to Cohorts 1 and 2) [80]. The US NTRDA grading system includes categories 0-3 based on extent of disease upon radiological examination (Supplemental Figure 4). The categories ranged from 0 as the least extensive disease (i.e. least severe) to 3 with the most extensive. For these analyses, we combined categories 0 and 1 and categories 2 and 3 from the US NTRDA grading system. This was operationalized as a binary variable and the analysis was done using a logistic regression model that adjusted for sex and HIV status. We tested all SNPs with P<1×10^−5^ for the association with TBscore for association with CXR severity.

### eQTL analysis

For our eQTL analysis in the follow-up cohort of 72 subjects without active TB, we first generated a list of genes that were differentially expressed in response to MTB infection, so we could narrow the list to genes that are regulated in response to active infection. We first analyzed eQTL’s using the cross linear model that tests for interactions between genotype (coded additively) and MTB stimulation status and its association with expression in a linear regression model. For our cross-linear model, if Y=expression, X_1_=SNP(additive coding), X_2_=Age, X_3_=Sex, and X_4_ = MTB stimulation status, then the regression equation for this analysis was: Y= β_0_ +β_1_X_1_ + β_2_X_2_ + β_3_X_3_ + β_4_ (X_4_) + β_5_ (X_1_X_4_) + ε. We followed up eQTL’s that showed an FDR < 0.1 for the interaction term by testing them for association with gene expression in an analysis that stratified by MTB stimulation. All models were adjusted for age and sex. To meet the definition of a stimulation dependent eQTL, a SNP had to show a statistically significant effect (an FDR < 0.1) in the MTB stimulated samples and a non-significant effect in the unstimulated samples (i.e., it had to be active specifically within the context of active MTB infection). We examined our list of severity-associated SNPs (i.e. those with P<1×10^−5^ for association with TBscore) within this list of genes that were differentially expressed in response to MTB stimulation (e.g., genes with significant interaction terms in the model above). All eQTL analysis was performed using the Matrix eQTL package in R v3.6.3[81]. Additional details about the eQTL analysis are found in the Supplemental Methods. Thus, the primary goal was to identify eQTL’s solely active after MTB stimulation, so we could narrow our results down to variants that play a role within the context of active TB that influenced severity. To determine if the eQTL effects observed in the whole cohort differ between those who are RSTR or LTBI, we performed a stratified analysis among only LTBI subjects and observed how this affected the associations we identified (eg. first order term).

In addition to the eQTL analysis using the data from our monocyte derived macrophage samples, we used FUMA GWAS to query a number of publicly available eQTL databases with the severity associated SNPs [78]. FUMA simultaneously queried several databases to see if these SNPs are eQTL’s for any of the genes and in any of the tissues included in the eQTL catalogue, eQTLgen, BIOSQTL, Blood eQTL Browser, DICE, xQTL Server, and GTEx v8 databases [59, 82-87]. In order to enrich these results and look for common biological functions, we uploaded the list of genes for which these SNPs were eQTL’s into the STRING database[88]. This database can implicate protein-protein interactions (PPIs) between the proteins downstream of the genes of interest as well as look for pathway enrichment such as KEGG or Gene Ontologies. PPIs among genes for which the SNPs from our GWAS analysis showed evidence of regulation can link their changes in expression to potential functional roles for their associated proteins and help explain why SNPS associated with severity are regulating these genes.

## Data Availability

All summary data is within the manuscript or supporting material. Because of the IRB restriction on the data from Uganda, individual level data are only available upon request from the Uganda Genetics of TB Data Access Committee by contacting the committee chair Dr. Sudha Iyengar.

## Acknowledgements

We would like to acknowledge the invaluable contribution made by the study medical officers, health visitors, laboratory and data personnel: Dr. Lorna Nshuti, Dr. Roy Mugerwa, Dr. Sarah Zalwango, Dr. Christopher Whalen, Dr. Deo Mulindwa, Dr. Christina Lancioni, Allan Chiunda, Denise Johnson, Bonnie Thiel, Mark Breda, Dennis Dobbs, Hussein Kisingo, Mary Rutaro, Albert Muganda, Richard Bamuhimbisa, Yusuf Mulumba, Deborah Nsamba, Barbara Kyeyune, Faith Kintu, Mary Nsereko, Gladys Mpalanyi, Janet Mukose, Grace Tumusiime, Pierre Peters, Annet Kawuma, Saidah Menya, Joan Nassuna, Alphonse Okwera, Keith Chervenak, Karen Morgan, Alfred Etwom, Micheal Angel Mugerwa, and Lisa Kucharski. We would like to acknowledge and thank Dr. Francis Adatu Engwau, Head of the Uganda National Tuberculosis and Leprosy Program, for his support of this project. We would like to acknowledge the medical officers, nurses and counselors at the National Tuberculosis Treatment Centre, Mulago Hospital, the Ugandan National Tuberculosis and Leprosy Program and the Uganda Tuberculosis Investigation Bacteriological Unit, Wandegeya, for their contributions to this study. This study would not be possible without the generous participation of the Ugandan patients and families.

## Author Contributions

MLM developed the hypotheses, conducted the analyses, drafted the manuscript, and finalized the manuscript. JS and TRH generated the RNA-seq data and edited the manuscript. HH conducted the original eQTL analyses and approved the final manuscript. LLM curated the clinical dataset and edited the manuscript. H M-K and WHB oversaw the clinical study and approved the final manuscript. WSB advised on bioinformatic analyses and approved the final manuscript. SMW developed hypotheses, supervised the analyses, drafted the manuscript and finalized the manuscript. CMS oversaw the clinical study, developed hypotheses, supervised the analyses, drafted the manuscript, and finalized the manuscript.

### Appendices

#### Supplemental Methods

##### Supplemental Data File

**Supplemental Table 1. Components of the TBscore and Points Contributed to Final Score**

**Supplemental Table 2. Studies of eQTL’s and Gene Expression**

**Supplemental Table 3. Table of all SNPs showing significant association with TBscore, accompanied by Summary Statistics from CXR GWAS Analysis** (excel file)

**Supplemental Table 4. Table of all SNPs showing P<0.05 and Same Direction of Effect for Association with TBscore in Both Cohorts, accompanied by Meta-Analytic Statistics** (excel file)

**Supplemental Table 5. MAF, P-value, and β value for rs1848553 Across Cohorts 1 and 2**

**Supplemental Table 6. Allele Frequencies for rs1848553 in 1000G Project in African Populations and Non-African Super-Populations**

**Supplemental Table 7. Association with TBscore for SNPs near Genes Previously Associated with TB Susceptibility**

**Supplemental Table 8. Association with TBscore for SNPs within +/- 50Kb of *IL12B*** (excel file)

**Supplemental Table 9. Cohort Characteristics for Ugandan Subjects in Matrix eQTL Analysis**

**Supplemental Table 10. Table of all Differentially Expressed Genes** (excel file)

**Supplemental Table 11. Table of all eQTL’s from SNPs Associated with TBscore at P<1e-05 (**excel file)

**Supplemental Table 12. rs2976562 Allele Frequencies in 1000G Project**

**Supplemental Figure 1. Imputation and QC of SNPs in Cohorts 1 and 2**

**Supplemental Figure 2. Plot of PC1 vs. PC2 in Cohort 1**

**Supplemental Figure 3. Plot of PC1 vs. PC2 in Cohort 2**

**Supplemental Figure 4. Categories for X-Ray severity**

**Supplemental Figure 5. Quantile-Quantile Plot for P-values for Association between SNPs and TBscore in Cohort 1**

**Supplemental Figure 6. Quantile-Quantile Plot for P-Values for Association Between SNPs and TBscore in Cohort 2**

**Supplemental Figure 7. Manhattan Plot of P-values for association between SNPs and TBscore in Cohort 1**

**Supplemental Figure 8. Manhattan Plot of P-values for association between SNPs and TBscore in Cohort 2**

**Supplemental Figure 9. Manhattan Plot for Meta-Analytic P-values for association between SNPs and CXR Extent**

**Supplemental Figure 10. Q-Q Plot for Meta-Analytic P-Values for association between SNPs and CXR Extent**

**Supplemental Figure 11. Decile Regression between rs184553 and TBscore, adjusted for Sex and HIV**

**Supplemental Figure 12. Supplemental Figure 12. STRING Network for PPI’s from eQTL Response Genes**

**Supplemental Figure 13. Expression of *SLA* in Immune Cells from DICE Database**

